# The Impact of the Lab4 Probiotic on the Wellbeing of Healthcare Workers: An Exploratory, Double-Blind, Randomised, Placebo-Controlled Study

**DOI:** 10.64898/2026.09.29.26364268

**Authors:** D. Dokic, A.A. Jack, N. Coates, D.A. John, I. Garaiova, J.A.K. McDonald, J.R. Marchesi, D. Wang, S.F. Plummer, D.R. Michael

## Abstract

Healthcare workers face substantial occupational demands and exposure to respiratory infections during winter, but evidence that probiotics improve their health and wellbeing remains limited. This exploratory, double-blind, randomised, placebo-controlled trial assessed whether daily Lab4 supplementation affected upper respiratory tract infection (URTI) frequency and supported wellbeing in 104 healthcare workers. Daily diaries recorded symptoms defining URTI episodes, general-practitioner visits, illness-related work absence and antibiotic use. Wellbeing was assessed at baseline and weeks 8 and 16, and faecal microbiota were assessed using 16S rRNA gene sequencing. Participants receiving Lab4 reported better wellbeing than those receiving placebo. For probiotic versus placebo, the risk ratio (RR) for URTI occurrence was 0.60 (95% confidence interval [CI]: 0.30–1.20; p=0.1477), the incidence rate ratio (IRR) for episodes was 0.58 (95% CI: 0.27–1.23; p=0.1581), and the mean ratio for episode duration was 0.98 (95% CI: 0.72–1.32; p=0.8776). For general-practitioner visits, the RR for at least one visit was 0.48 (95% CI: 0.21–1.07; p=0.0737) and the IRR was 0.44 (95% CI: 0.18–1.06; p=0.0677). For illness-related work absence, the RR for at least one day absent was 0.75 (95% CI: 0.18–3.22; p=0.6991) and the IRR for absence days was 0.74 (95% CI: 0.16–3.32; p=0.6914). For antibiotic use, the RR for any use was 0.48 (95% CI: 0.20–1.18; p=0.1111) and the IRR for antibiotic-use days was 0.42 (95% CI: 0.16–1.12; p=0.0820). No significant between-group differences in gut microbiota were detected in the subset providing faecal samples. Illness-related estimates generally favoured Lab4, although between-group differences were not statistically significant. These exploratory findings support evaluation in a larger, prospectively powered trial.

## 1. Introduction

The gut microbiota comprises the community of microorganisms inhabiting the gastrointestinal tract and contributes to human health through nutrient metabolism, maintenance of intestinal barrier integrity, immune regulation and communication along the gut–brain axis **(1–3)**. The gut microbiota may also influence susceptibility and responses to respiratory infection through the gut–lung axis **(4)**. Gut microbial metabolites have been shown to enhance antiviral immunity in models of respiratory syncytial virus infection **(5)**. Through these effects on metabolic, immune, neurological and respiratory health, the gut microbiota contributes to the overall wellbeing of the host.

Wellbeing encompasses physical and emotional health, the ability to function effectively and to participate fully in society **(6,7)**. Even among generally healthy individuals, the demands of everyday life, including occupational stress, poor diet and unhealthy lifestyle behaviours, can adversely affect overall wellbeing **(8,9)**. Wellbeing is also influenced by seasonality**(10)**; during the winter months, respiratory infections, particularly upper respiratory tract infections (URTIs), are more common, while rates of workplace absenteeism, healthcare utilisation and antibiotic use increase **(11–13)**. Healthcare workers may be particularly vulnerable during this period because of the demands of a high-stress working environment and, among patient-facing staff, increased exposure to individuals with communicable illnesses **(14,15)**. These occupational and seasonal pressures highlight the need for approaches that help healthcare workers maintain their wellbeing during winter. Given the influence of the gut microbiota on physical and psychological wellbeing, modulation of the microbiota warrants investigation as one such approach.

Probiotics, defined as live microorganisms that confer a health benefit when administered in adequate amounts **(16)**, have been shown to influence the functionality and composition of the gut microbiota **(17)**, enhance resilience to respiratory tract and other infections **(18)** and improve mood and sleep quality **(19,20)**. However, findings vary with probiotic formulation, trial population and outcome assessed **(18,19)**. Only a few studies have assessed the effect of probiotic supplementation on respiratory tract infections in healthcare workers and report mixed results; a study with frontline medical staff during the COVID-19 pandemic found that *Streptococcus thermophilus* ENT-K12 reduced the incidence of respiratory tract infections **(21)**, whereas a placebo-controlled trial in nurses found no significant effect of *Lacticaseibacillus rhamnosus* HN001 on perceived stress or days with viral illness symptoms **(22)**.

The Lab4 probiotic consortium comprises *Lactobacillus acidophilus* CUL60 and CUL21, *Bifidobacterium bifidum* CUL20 and *Bifidobacterium animalis* subsp. *lactis* CUL34. Previous trials provide a rationale for investigating its effects on the wellbeing of healthcare workers; daily supplementation with Lab4 has been found to provide wellbeing-related benefits in other populations **(23)**, has been shown to reduce URTI symptoms in adults **(24)** and children **(25)**, and modulate the composition and functionality of the gut microbiota **(26,27)**. *In vitro* work with the Lab4 consortium has shown its capability to enhance antibacterial and antiviral immune responses **(28)**.

In this exploratory, double-blind, randomised, placebo-controlled trial, we investigated whether daily supplementation with Lab4 for 16 weeks could support the wellbeing of healthcare workers during winter. We hypothesised that supplementation would be associated with a lower burden of respiratory illness and improved self-reported wellbeing compared with a placebo. Illness-related outcomes included the occurrence, incidence and duration of symptom-defined URTI episodes, general-practitioner visits, illness-related work absence and antibiotic use. The Wellbeing questionnaire assessed general wellness, perceived health, mood, energy, and sleep quality, providing participant-reported indicators of selected aspects of wellbeing. Faecal microbiota composition was also examined to explore microbial responses to supplementation. The study sought to identify signals of potential benefit across these measures.

### 2. Materials and Methods

### 2.1. Study Approval

This study was conducted in accordance with the ethical principles of the Declaration of Helsinki and was approved by the Ethics Committee for Clinical and Other Investigations related to Medicines and Medical Supplies within the Agency for Medicines and Medical Supplies of the Republic of Macedonia (Submission reference: 11-372/1 Approval date 20/02/2017) and registered with the ISRCTN registry (ISRCTN90136957 Registration date: 02/04/2020). Registration occurred after participants had completed the trial but before the data was analysed. The principle investigator was unaware that exploratory studies needed to be registered, so the registration was completed retrospectively.

### 2.2. Study Design

This exploratory, 16-week, double-blind, randomised, placebo-controlled trial was conducted at the University Clinic of Pulmonology and Allergology, Skopje, North Macedonia, with equal allocation between two study groups.

### 2.3. Recruitment and Randomisation

Adults aged 18-70 years old working in a healthcare setting (hospitals, surgeries, pharmacies) but not necessarily patient-facing, were recruited during the winter period. Eligibility criteria included ability to provide written informed consent, no probiotics consumed within the previous 2 weeks, nor antibiotics within the previous 3 weeks of the trial start, not pregnant or lactating, willing to refrain from taking products/medications that stimulate immune function/inflammation and willing to maintain normal diet and lifestyle.

Participants were sequentially allocated to one of 2 study arms of the study in a 1:1 ratio according to a computer-generated random sequence using permuted block randomisation with a block size of four (SAS PROC PLAN (SAS v9.4)). The study intervention was randomised at the point of manufacture, prior to its arrival at the trial site. The allocation sequence was not available to any member of the research team until databases had been completed and locked but was held at the trial site in tamper-proof sealed envelopes in case of emergency.

### 2.4. Study Product

The probiotic capsules contained 50 billion colony-forming units (CFU) of *Lactobacillus acidophilus* CUL60, *Lactobacillus acidophilus* CUL21, *Bifidobacterium bifidum* CUL20, and *Bifidobacterium animalis* subsp. *lactis* CUL34 on a microcrystalline cellulose carrier. The matched placebo product was microcrystalline cellulose and maltodextrin and was similar in appearance to the active product. All products were recommended to be stored at 4-8°C.

### 2.5. Intervention

Participants were asked to consume one capsule daily with food (with or without a cool drink) and to avoid consumption within 2 hours of any antibiotic intake. Participants were provided with pots of product at baseline and at the week 8 trial centre visit. Unused capsules were recorded for compliance monitoring and viability testing.

### 2.6. Study Aims

The study explored the hypotheses that compared to a placebo; 16 weeks of Lab4 supplementation would be associated with a lower URTI burden, fewer visits to general practitioners (GPs), less antibiotic usage, less illness-related work absence, improved wellbeing, and differences in the faecal microbiota. The health-related outcomes were: the occurrence, incidence rate, duration and time to first URTI episode, general-practitioner visits, illness-related work absence, and antibiotic use, measured with symptom diaries. Wellbeing was assessed using a five-item questionnaire and composite score. Faecal bacterial diversity, community composition, and genus-level abundance were assessed.

### 2.7. Data and Sample Collection

The sequence of data and sample collection during the study is shown in **Fig 1A**. Symptom diaries were completed daily; wellbeing was assessed at baseline, week 8 and week 16; faecal samples were provided voluntarily by participants within 48 hours of the baseline and week 16 visits. Harms were assessed non-systematically and any spontaneously reported events were documented.

**Fig 1.**
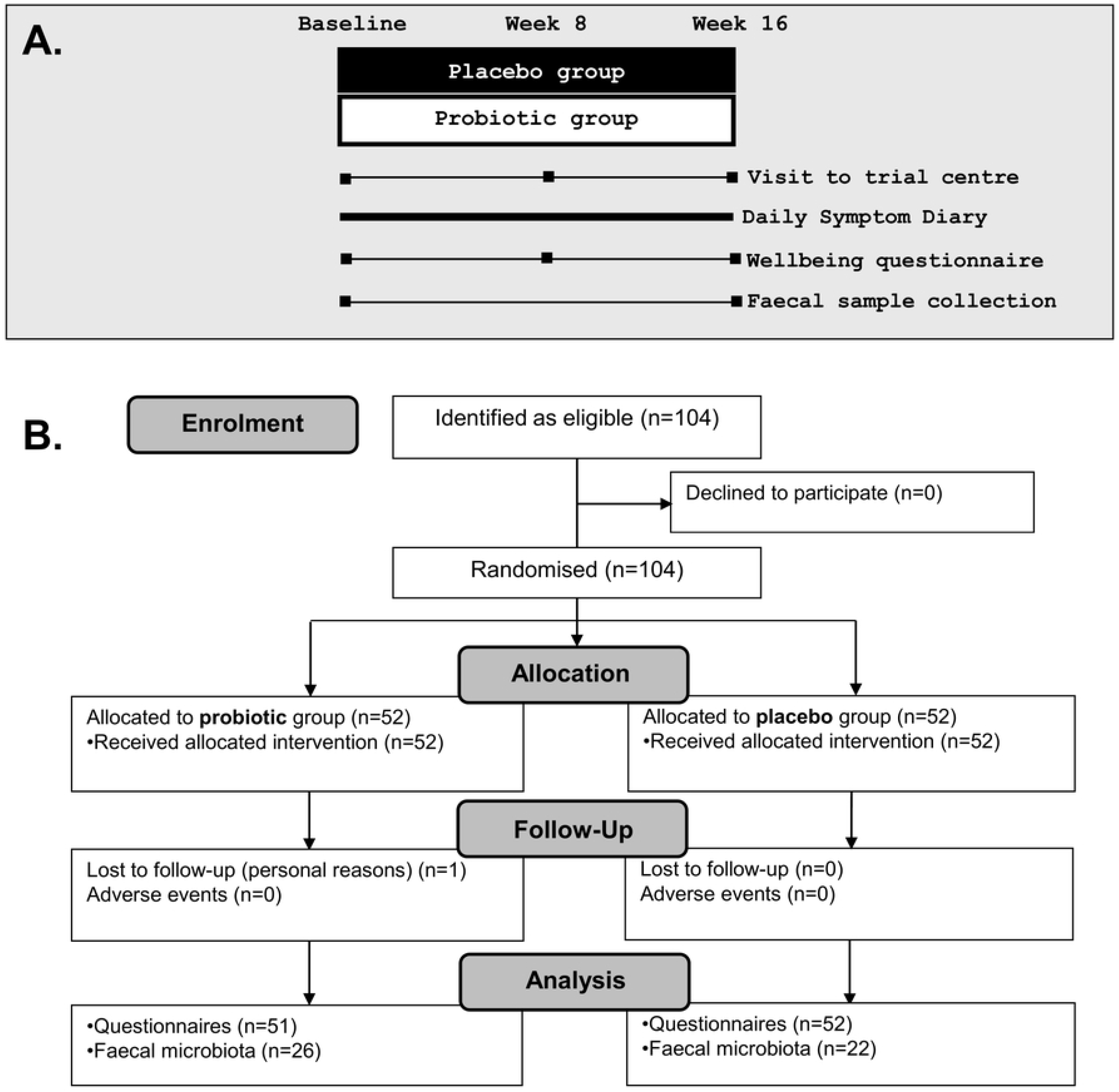
Participant flow diagram (A) and study design (B).

### 2.8. Daily Symptom Diaries

The symptom diaries monitored *chills, sneezing, coughing, blocked nose, runny nose, headaches, sore throats* and *malaise* according to Jackson *et al* (29). Each symptom was scored as 0 (none or mild), 1 (moderate) or 2 (severe), giving a daily total score from 0 to 16. A URTI episode was defined as a daily total score of at least 5 on each of three or more consecutive days, and distinct episodes were separated by at least two symptom-free days. Visits to GPs, absenteeism from work due to illness and antibiotic use were also recorded.

### 2.9. Wellbeing

A 5-item 1-week recall questionnaire(30) used previously (23,24,30), recorded *General Wellness, State of Mood, State of Health, State of Energy and Sleep Quality*) on a 10-point Likert scale from 1 (very poor) to 10 (very good). Wellbeing Score was calculated from the sum of the five items, multiplied by 2 and expressed as a percentage, with worst and best possible scores of 10% and 100%, respectively.

### 2.10. Faecal Sample Collection

Participants providing faecal samples used a Fe-col collection kit (Alpha Laboratories, Hampshire, UK) and transported the samples in 30 ml Universal Containers (SLS, UK). Samples were stored anaerobically at −80 °C pending analysis. Participants collecting samples at home were asked to refrigerate samples and return them to the trial centre within 48h of collection.

### 2.11. DNA Extraction and 16S rRNA Gene Sequencing

DNA was extracted from faeces using the QIAamp^®^ Fast DNA Stool Mini Kit (Qiagen, Germany). Samples were homogenised in Matrix Lysing D tubes (MPBIO) and a FastPrep^®^-24 bead beater (MPBIO, United States). Eluted genomic DNA was quantified using a Qubit^®^ (Thermo Fisher Scientific, United States), normalised to 5 ng in tris-acetate-ethylenediaminetetraacetic acid (TAE) and stored at −20 °C. Sample libraries were prepared following Illumina’s 16S Metagenomic Sequencing Library Preparation Protocol with the following modifications. Bifidobacteria sequences (31) were amplified in addition to the V1–V2 regions of the 16S rRNA gene. The index PCR products were cleaned up and normalised using the SequalPrep Normalization Plate Kit (Life Technologies, Paisley, United Kingdom). Sample libraries were quantified using the NEBNext Library Quant Kit for Illumina (New England Biolabs, Hitchin, United Kingdom). Sequencing was performed on an Illumina MiSeq platform (Illumina Inc., Saffron Walden, United Kingdom) using the MiSeq Reagent Kit v3 (Illumina) and paired-end 300 base pair (bp) chemistry. Negative controls for the sequencing were reactions containing no template DNA. Paired-end 16S rRNA gene sequencing reads were trimmed, chimeric sequences were removed, and paired ends were merged using DADA2 (32) to obtain amplicon sequence variants (ASVs) which were aligned against the SILVA database v138.1 to assign taxonomy (33).

### 2.12. Statistical Analyses

Statistical analyses were conducted on randomised participants who provided post-baseline data. Missing data was not imputed. All statistical analyses were performed in R (version 4.4.3) unless otherwise stated. All models included group, age and BMI as fixed effects. Models for outcomes with repeated measurements also included a participant-level random intercept. Model diagnostics were assessed using simulated residuals from DHARMa (v0.4.7), and estimated marginal means (EMMs) were determined using emmeans (version 2.0.3) (34). Given the exploratory nature of this trial, we present p-values without correction for multiple comparisons to minimise the risk of Type II errors in a small study population. Values of p ≤ 0.05 were considered statistically significant.

The proportions of participants reporting a URTI episode, absence from work, GP visit or antibiotic use during the study period were analysed using binomial generalised linear models (GLM) with logit link. Model-adjusted probabilities were obtained by back-transforming the EMMs from the log-odds scale to the probability scale. The risk ratio (RR) is the adjusted probability in the probiotic group divided by the probability in the placebo group; a risk ratio < 1 indicated a lower probability of an occurrence in the probiotic group, whereas a risk ratio > 1 indicated a higher probability.

The incidence rates of URTI episodes or days reporting absence from work, GP visit(s) or antibiotic use during the study period were analysed using a GLM with a log-link negative binomial family and an offset for the logarithm of the number of days providing data. Model-adjusted incidence rates were expressed per 100 participant-days. Incidence rate ratios (IRRs) are the adjusted incidence rate in the probiotic group divided by that in the placebo group. An IRR < 1 indicates a lower rate in the probiotic group, whereas an IRR > 1 indicated a higher rate.

The duration of individual URTI episodes was analysed using a log-link Poisson generalised linear mixed model (GLMM) with a participant random intercept to compare the mean length (days) of URTI episodes between the groups, while accounting for repeated testing of participants. Time-to-first URTI episode was analysed with the Kaplan-Meier method with the log-rank Mantel-Cox test (GraphPad Prism, V10.6.1).

Wellbeing Score was analysed using a covariate adjusted linear mixed model (LMM) with a participant-level random intercept and group, time-point and a group-by-time point interaction as additional fixed effects.

Individual Wellbeing items were analysed using cumulative link mixed models (CLMMs, *ordinal (version 2025.12.29)* (35)) to estimate the log-odds of the probiotic group reporting a higher score than the placebo group at week 8 and week 16. The proportional odds assumption was verified for each outcome using nominal and scale tests on equivalent cumulative link models without the participant-level random intercept. Adjacent scores with few observations were pooled together to stabilise models and ensure test assumptions were met. Group estimated marginal means were obtained on the latent model scale, and contrasts were exponentiated and reported as odds ratios (OR) with 95% confidence intervals representing the odds of reporting a higher score for the probiotic group/placebo group. An OR > 1 indicates that probiotic participants were more likely to report a higher score than placebo participants, whereas an OR < 1 indicates they were less likely. As a sensitivity analysis, Mann–Whitney U tests were used to compare the score distributions between groups separately at weeks 8 and 16.

### 2.13. Statistical Analysis of Faecal Microbiota

Bacterial community analyses were performed in R using phyloseq, vegan, and ggplot2 (36,37). Samples containing >1,000 sequencing reads were retained for analysis and no abundance/prevalence filtering was applied. All faecal microbiota models included group, age, BMI and sample sequencing depth as fixed effects. Group differences in week 16 genus-level Shannon and Simpson alpha diversities were assessed using ANCOVA with baseline score as an additional fixed effect. Genus-level beta diversity was evaluated using robust Aitchison distance and compared at baseline and week 16 using covariate adjusted PERMANOVA (with 9,999 permutations) and betadisper/permutest (9,999 permutations). Robust Aitchison distances were visualised using principal coordinate analysis. MaAsLin3 (38) was used to identify differentially abundant genera associated with the group-by-time point interaction. No prevalence or abundance filter was applied and a total of 208 genera were analysed. Counts were total-sum scaled and log transformed for analysis.

### 2.14. Use of Generative Artificial Intelligence

ChatGPT 5.6 Sol was used for coding assistance in R and language improvement. Generative AI was not used for idea generation or exploration.

## 3. Results

### 3.1. Participant Flow and Baseline Characteristics

Participants were recruited between 5/12/2017 to 22/12/2017. A total of 104 participants were randomised and 103 were included in the reported analyses (**Fig 1B**). One participant dropped out of the probiotic group for personal reasons and there were no reported serious adverse events. Compliance to product exceeded 98% in both groups. The baseline characteristics of the study participants are presented in **Table 1**.

**Table 1.**
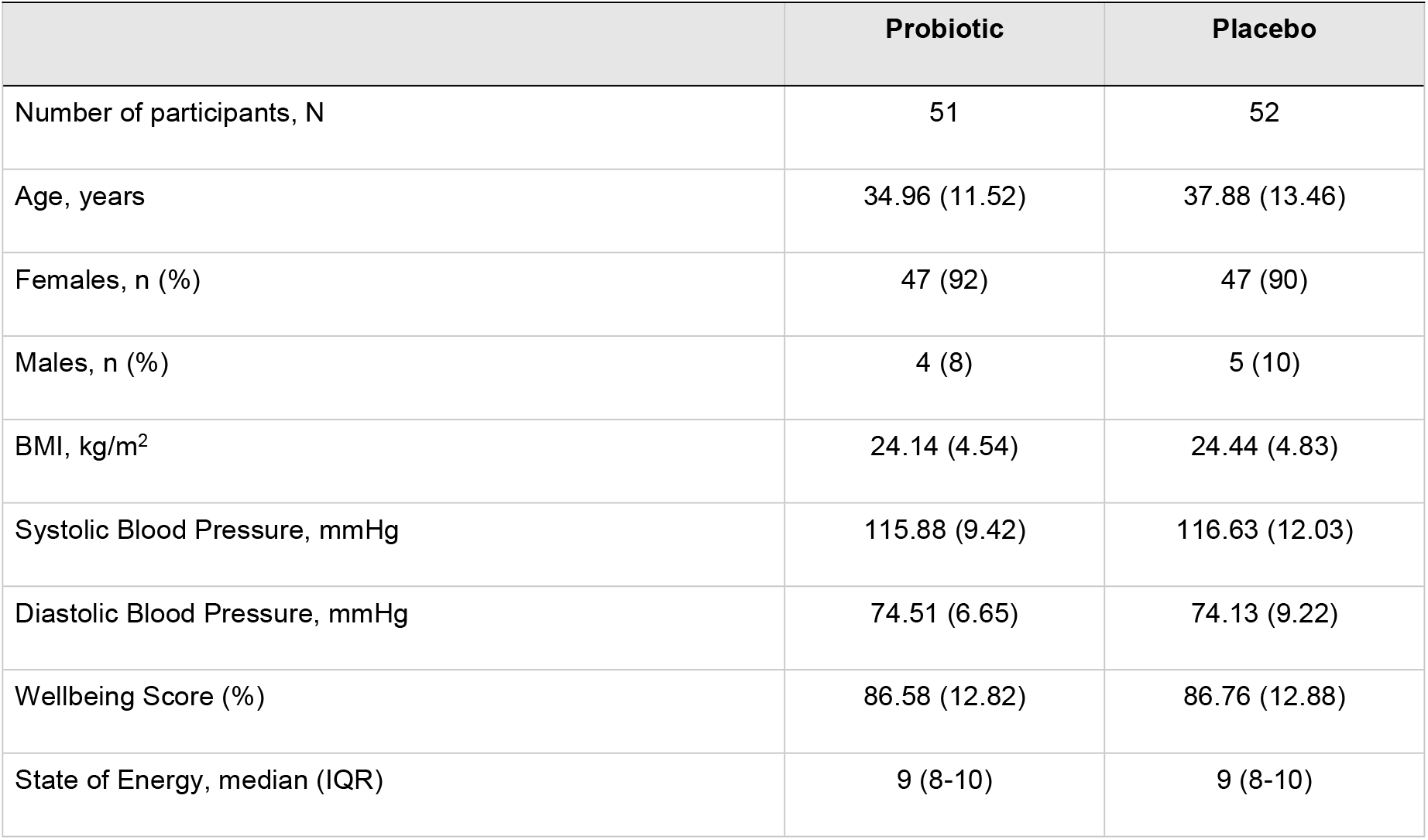
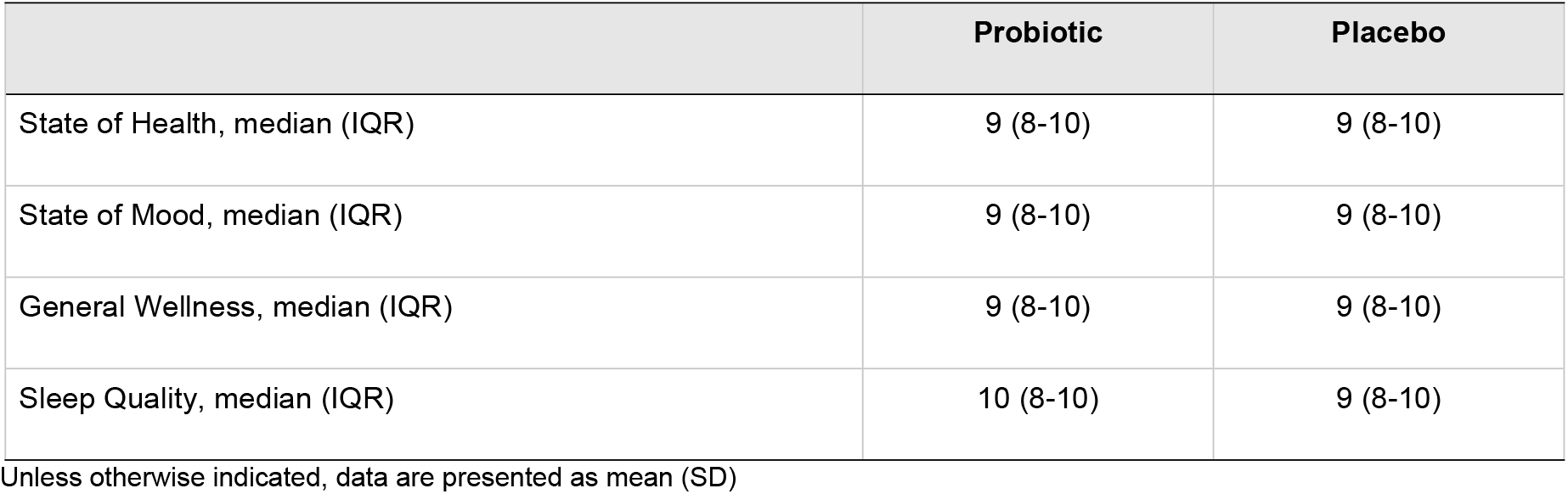
Baseline characteristics.

### 3.2. Upper Respiratory Tract Infection (URTI)

At least one URTI episode was reported by 10/51 (19.6%) participants in the probiotic group and 17/52 (32.7%) in the placebo group (RR: 0.60; 95% CI: 0.30 to 1.20; *p=*0.1477; **Table 2**). The Kaplan–Meier curves for time to first URTI episode show separation between the groups, with a greater proportion of participants in the probiotic group remaining URTI-free (log-rank test: *p=*0.1100; **Fig 2**). A total of 14 URTI episodes were reported in the probiotic group, compared with 23 in the placebo group (IRR: 0.58; 95% CI: 0.27 to 1.23; *p=*0.1581). The adjusted mean duration of a URTI episode was 5.55 days in the probiotic group and 5.68 days in the placebo group (mean ratio: 0.98; 95% CI: 0.72 to 1.32; *p=*0.8776).

**Table 2.**
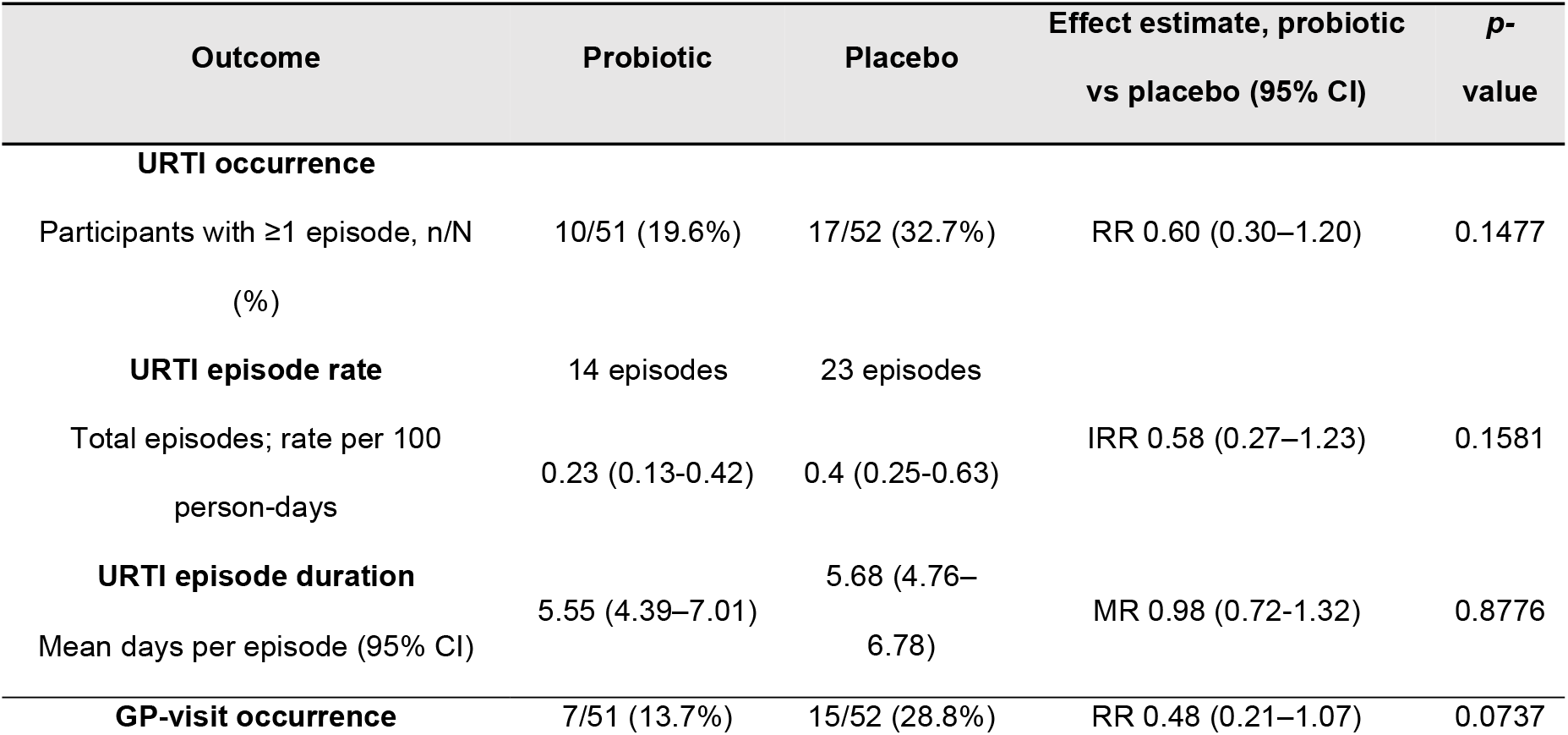
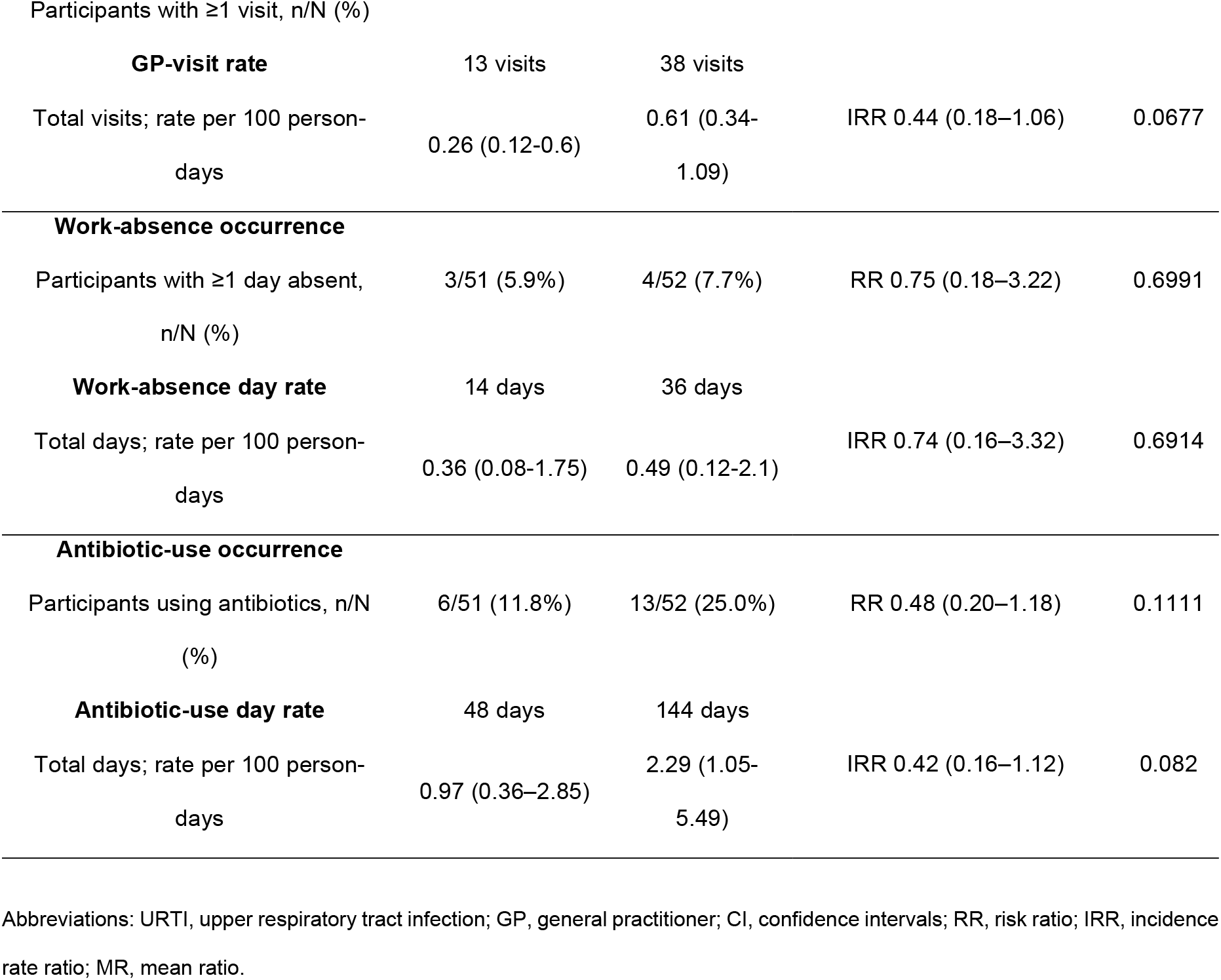
URTI, GP visits, absenteeism and antibiotic use.

**Fig 2.**
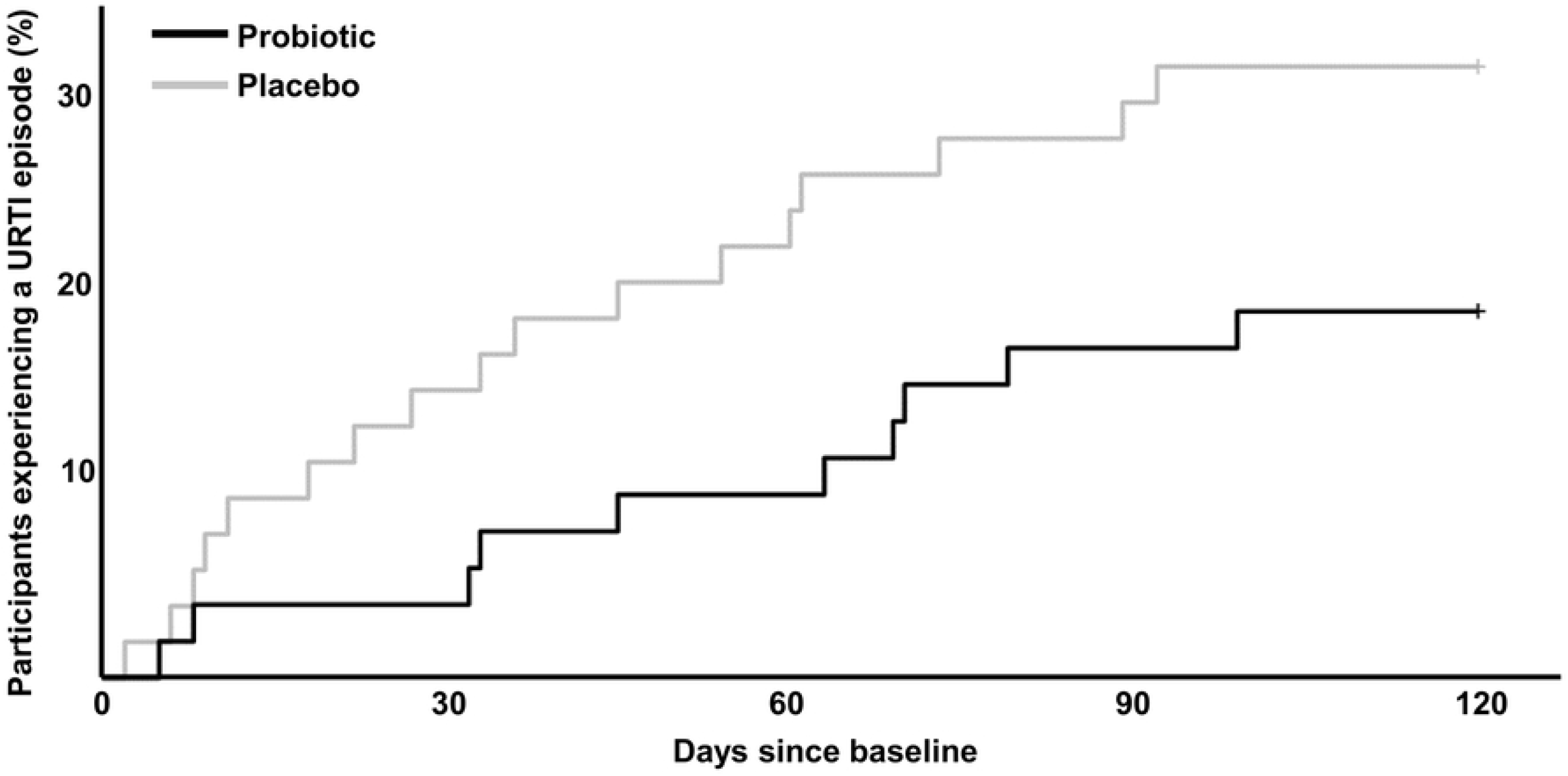
Time to first URTI episode.

### 3.3. General Practitioner Visits

At least one GP visit was reported by 7/51 (13.7%) participants in the probiotic group and 15/52 (28.8%) in the placebo group (RR: 0.48; 95% CI: 0.21 to 1.07; *p=*0.0737; **Table 2**). A total of 13 GP visits were reported in the probiotic group and 38 in the placebo group (IRR: 0.44; 95% CI: 0.18 to 1.06; *p=*0.0677).

### 3.4. Absence from Work

Illness-related absence was reported by 3/51 (5.9%) participants in the probiotic group and 4/52 (7.7%) in the placebo group (RR: 0.75; 95% CI: 0.18 to 3.22; *p=*0.6991). A total of 14 absence days were reported in the probiotic group and 36 in the placebo group (IRR: 0.74; 95% CI: 0.16 to 3.32; *p=*0.6914).

### 3.5. Antibiotic Use

Antibiotic use was reported by 6/51 (11.8%) participants in the probiotic group and 13/52 (25.0%) in the placebo group (RR: 0.48; 95% CI: 0.20 to 1.18; *p=*0.1111). The groups reported 48 and 144 antibiotic-use days, respectively (IRR: 0.42; 95% CI: 0.16 to 1.12; *p=*0.0820). Details of antibiotic classes used are reported in **Supplementary Table S1**.

### 3.6. Wellbeing

The Wellbeing Score increased in both groups; in the probiotic group from 86.58% at baseline to 91.06% at week 8 (adjusted mean change of 4.48%; 95% CI: 3.16% to 5.78%, *p<*0.0001) and 94.28% at week 16 (7.70%; 95% CI: 5.06% to 10.32%, *p<*0.0001), and from 86.76% at baseline to 88.74% (1.98%; 95% CI: 0.72% to 3.2%, *p=*0.0068) and 89.88% (3.12%; 95% CI: 1.4% to 4.84%, *p<*0.0001) in the placebo group. The between-group difference in Wellbeing Score was 2.58 percentage points (95% CI: 0.58 to 4.56; *p=*0.0111) at week 8 and 4.64 percentage points (95% CI: 2.66 to 6.62; *p<*0.0001) at week 16 (**Fig 3**). For the individual Wellbeing items, there were no between-group differences at Week 8 (all *p≥*0.2601; details in **Supplementary Table S2**), but at Week 16, the probiotic group had greater odds of reporting higher scores for State of Energy (OR 36.30, 95% CI 3.60–365.80; *p=*0.0023), State of Health (OR 22.02, 95% CI 1.47–330.25; *p=*0.0252), and State of Mood (OR 11.25, 95% CI 1.32–95.73; *p=*0.0268). No differences were detected for General Wellness or Sleep Quality at Week 16 (*p=*0.3767 and *p=*0.7140, respectively).

**Fig 3.**
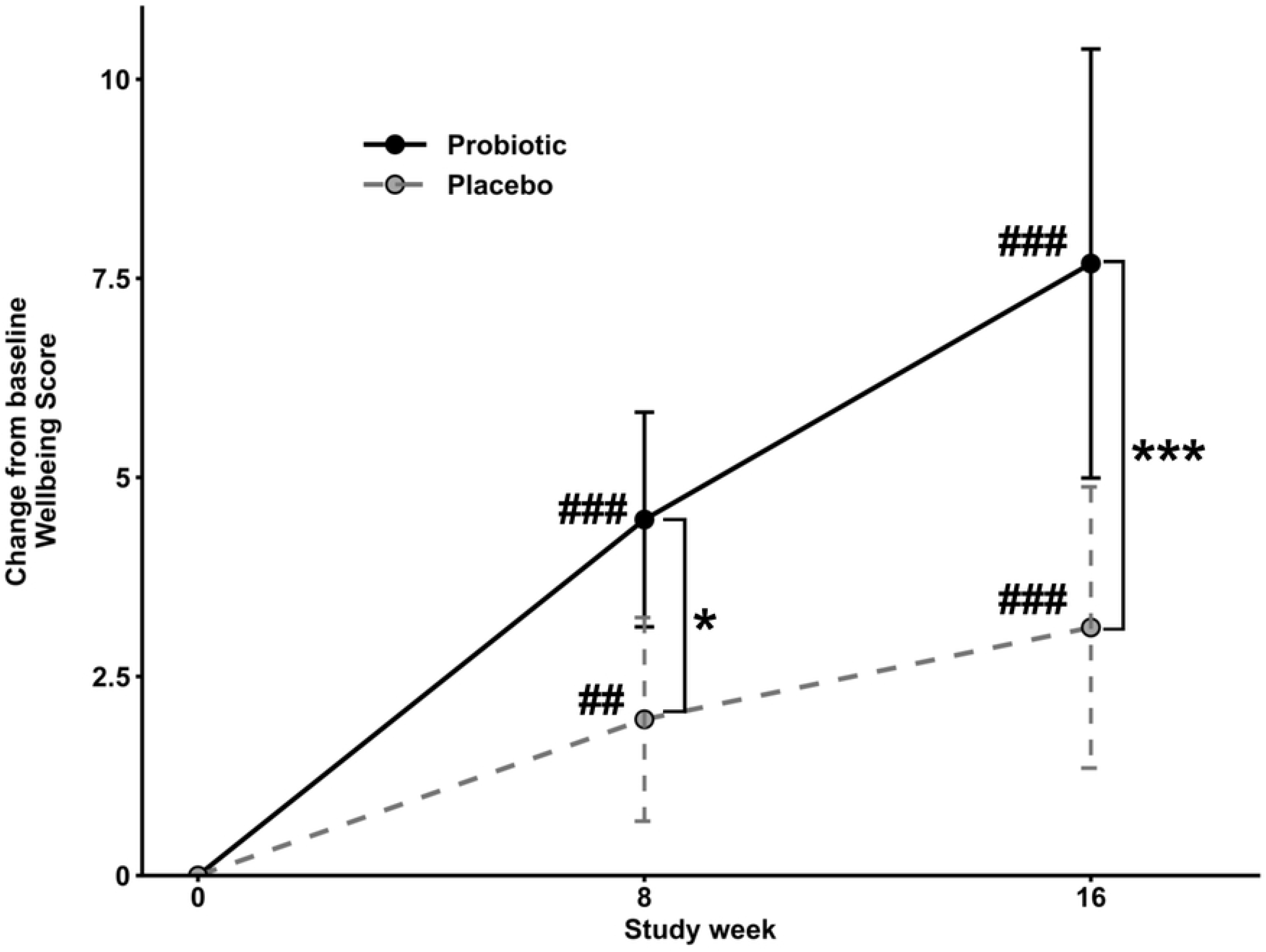
Changes in Wellbeing Score during the study. Data are presented as mean change from baseline with 95% confidence intervals. Between-group comparisons: *p ≤ 0.05 and ***p ≤ 0.001; within-group comparisons versus baseline: ##p ≤ 0.01 and ###p ≤ 0.001.

### 3.7. Faecal Microbiota Composition

In this study, faecal samples were provided voluntarily by 48 participants (probiotic n=26, placebo n=22). The baseline characteristics of the population providing the samples were representative of the total study population (**Supplementary Table S3**).

At week 16 there were no significant between-group differences in Shannon (adjusted mean difference of -0.07; 95% CI: -0.21 to 0.08, *p=*0.3747; **Fig 4A**) or Simpson alpha diversity (-0.007; 95% CI: -0.029 to 0.014, *p=*0.4877). Beta diversity was comparable between groups at baseline (PERMANOVA group variable: R^2^ = 0.006, F = 0.311, *p=*0.7850; dispersion *p=*0.8791) but at week 16 there indications of compositional differences (R^2^ = 0.046, F = 2.27, *p=*0.0888; dispersion *p=*0.5412; **Fig 4B**). Differential abundance analysis identified a number of bacterial genera that were associated with probiotic supplementation (**Fig 4C**): *Fusicatenibacter* (*p=*0.0241), *Peptococcus* (*p=*0.0002), *Lachnospiraceae* UCG-010 (*p=*0.0400), *Parasutterella* (*p=*0.0145) and *Eisenbergiella* (*p=*0.0463) were positively associated and *Colidextribacter* (*p=*0.0221) was negatively associated with probiotic consumption. Although none of these genera remained significant after false discovery rate adjustment, we report these nominal associations in keeping with the exploratory, signal-generating nature of the study, as they provide taxonomic context for the study outcomes.

**Fig 4.**
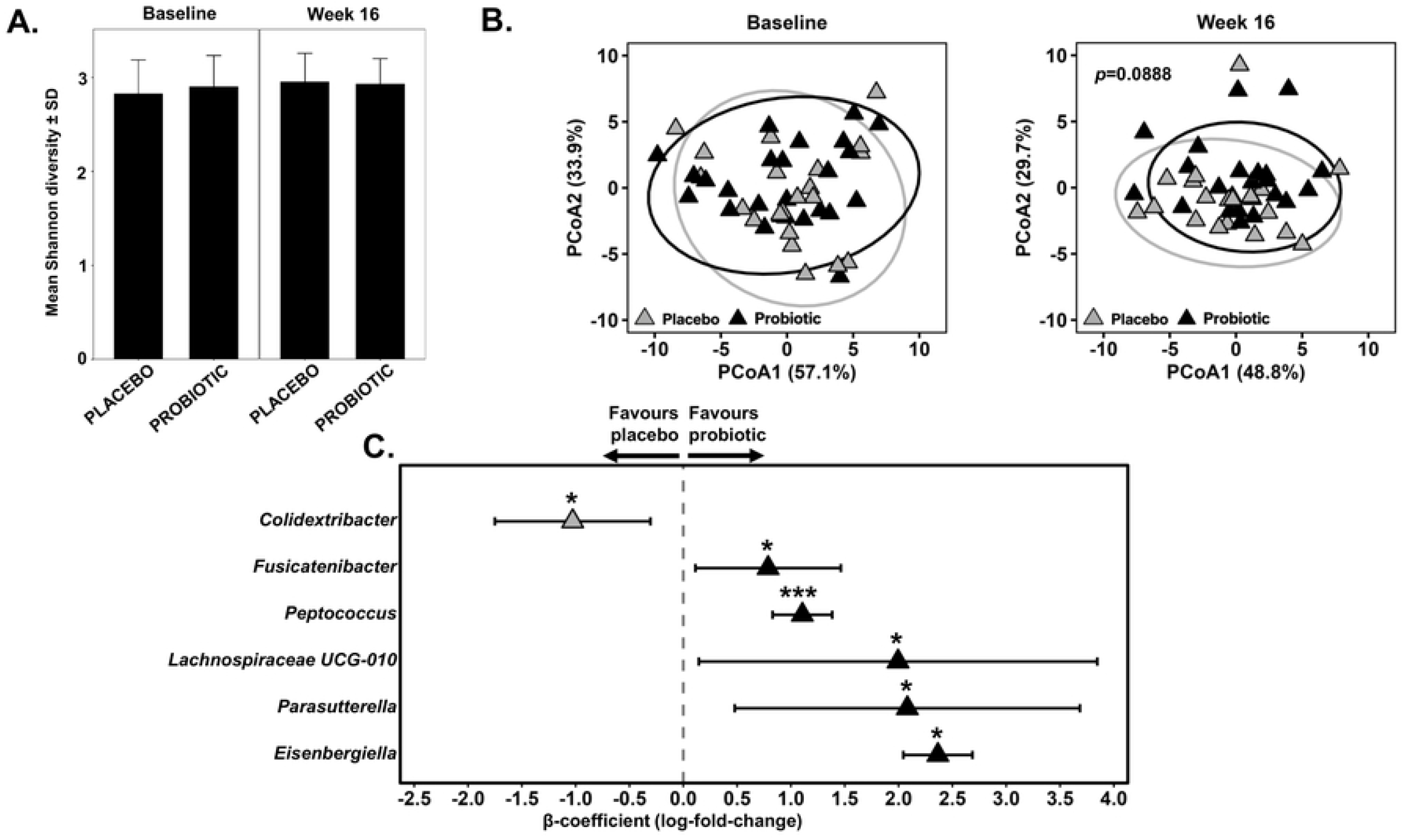
The composition of the faecal microbiota. (A) Alpha diversity using the Shannon index. (B) Beta diversity by principal coordinate analysis of robust Aitchison distances. (C) Differential abundance of genera (probiotic vs. placebo); positive β-coefficients indicate positive correlation wsith probiotic supplementation, whereas negative β-coefficients indicate a negative correlation.

## 4. Discussion

In this exploratory, randomised, double-blind, placebo-controlled study, participants supplemented with the Lab4 probiotic consortium for 16 weeks showed greater wellbeing than those receiving a placebo. No statistically significant between-group differences were detected in URTI burden, general practitioner visits, illness-related absence or antibiotic use. However, the estimates generally favoured the probiotic group, which reported fewer URTI episodes and fewer days with URTIs, GP visits, and antibiotic use, as well as lower proportions of participants reporting these outcomes. Notwithstanding, the confidence intervals indicate imprecision, which likely reflect the small sample size and low number of events in this exploratory trial.

Evidence for probiotic prevention of respiratory illness is mixed with some evidence of modest reductions in URTI duration or symptom burden (21,39–42), whereas other trials, including a study in nurses (22), have not detected a probiotic associated reduction in viral-illness days (18,43). Differences in probiotic strain, dose, population, season, and outcome definitions add to the heterogeneity between studies and limit direct comparisons. The direction of the estimates in this study is consistent with findings from previous Lab4 studies; in the PROMAGEN trial of overweight and obese adults there was a 40% reduction in the incidence of URTI-related symptoms (24), and in the PROCHILD studies, children receiving Lab4 had fewer occurrences and shorter durations of URTIs, as well as less antibiotic usage and fewer GP visits (25).

Baseline Wellbeing Scores were unexpectedly high, despite the occupational demands experienced by healthcare workers during the winter period, and exceeded scores reported in a study of NHS workers using the same questionnaire (44). Additionally, we did not see a decline in wellbeing over winter for the placebo group; in fact, participants reported improved wellbeing relative to baseline which might be due to an expectancy effect, regression to the mean, or the Hawthorne effect (45). Even so, wellbeing improved more in the Lab4 group, leading to between-group differences-a finding reported in previous studies with Lab4 (24), including in women with irritable bowel syndrome, where wellbeing improvements were associated with reductions in anxiety and/or depression (23). In contrast, healthy adults receiving a multistrain synbiotic for 6 weeks showed no changes in ‘State of Health’ or ‘State of Energy’ which was attributed to the high baseline scores (30). A study of female healthcare workers receiving *Lactobacillus delbrueckii* ssp. *bulgaricus* OLL1073R-1 showed improved general health and vitality (46).

In this study, Lab4 supplementation did not produce robust changes in the gut microbiota, but there were indications of altered community composition at week 16. The enrichment of *Fusicatenibacter* and *Lachnospiraceae UCG-010* is of interest because reduced abundances of *Fusicatenibacter* and *Lachnospiraceae* have been reported in individuals with respiratory tract infections (47). Changes to the gut microbiota have previously been reported in healthy adults receiving Lab4 (27) so the lack of detectable effect in this study may reflect the low number of participants who provided stool samples. Gut microbiota modulation is sometimes reported in other probiotic studies (17).

This study has several strengths: a diversity of measures were included to provide a holistic assessment of the potential effect of the probiotic on the wellbeing of healthcare workers; it was run during the winter season when challenge from respiratory infections is higher; daily recording of symptoms minimised reliance on long-term recall. Also, a stringent definition for a URTI episode was applied, requiring both a minimum symptom burden and persistence over consecutive days to strengthen the outcomes. However, there are limitations that should also be considered. The study was exploratory, and the absence of a formal sample size calculation means findings should be interpreted as indicative and requiring confirmation in an adequately powered trial. URTI episodes were identified from self-reported, non-specific symptoms rather than from physician diagnosis or laboratory confirmation. The cohort was predominantly female, so sex-specific effects could not be estimated reliably. Finally, faecal samples were supplied voluntarily by fewer than half of randomised participants which further limited power to detect any microbiota differences.

### 5. Conclusions

In this exploratory trial, participants allocated to Lab4 reported greater improvements in perceived wellbeing than those receiving placebo at weeks 8 and 16. The Lab4 group reported a lower URTI burden, fewer general-practitioner visits and less antibiotic use, but between-group differences were not statistically significant. The observed signals support evaluation of Lab4 in a larger study of healthcare workers.

## Data Availability

The study protocol is available through the ISRCTN registration (ISRCTN90136957). The datasets underlying the findings reported in this study will be provided as Supporting Information files. Sequence data generated during the study are available from the European Nucleotide Archive (ENA) under accession number PRJEB122545.

## Acknowledgments

The authors thank Josh Kerry-Smith and Kathryn Loxley for technical assistance during the study.

## Supplementary Materials

The following supporting information accompanies this article: **Table S1, Breakdown of antibiotic usage during the intervention period; Table S2, Individual Wellbeing Item Analysis at Weeks 8 and 16; Table S3, Baseline characteristics of the participants providing faecal samples**.

## Notes

### Competing Interest Statement

Competing interests: D.R.M., A.A.J., N.C., D.A.J., I.G. and S.F.P. are or were employees of Cultech Ltd, which funded the study. These authors had no role in participant recruitment or data collection but contributed to study design, data analysis and interpretation, and/or manuscript preparation, review and approval. The remaining authors declare no conflicts of interest.

### Clinical Trial

ISRCTN90136957

### Author Declarations

The study was conducted in accordance with the ethical principles of the Declaration of Helsinki and was approved by the Ethics Committee for Clinical and Other Investigations related to Medicines and Medical Supplies within the Agency for Medicines and Medical Supplies of the Republic of Macedonia (submission reference 11-372/1 approved 20 February 2017). Written informed consent was obtained from all participants prior to participation in the study.

